# The Effect of Vitamin-D Supplementation on HDAC2 Levels in Stable COPD Patients

**DOI:** 10.64898/2026.04.05.26348641

**Authors:** Adyan Donastin, Danny Irawan, Effendy, Rifky Dwi Aditya Iryawan, Nuzlan Nuari, Betta Mega Oktaviana, Devy Yahya, Akbar Reza Muhammad

## Abstract

**Background:** Chronic Obstructive Pulmonary Disease (COPD) is the third leading cause of global mortality, with persistent lung inflammation contributing to disease progression. This inflammation is partly associated with reduced levels of histone deacetylase 2 (HDAC2). Previous studies suggest that Vitamin D may modulate HDAC2 levels. This study aimed to evaluate the effect of Vitamin D supplementation on HDAC2 expression in stable COPD patients. This experimental study aimed to evaluate the effect of vitamin D supplementation on HDAC2 expression in stable COPD patients at Jemursari Islamic Hospital.

**Methods:** Five COPD patients received a daily dose of 5000 IU of Vitamin D for three months. Serum levels of 25(OH)D3 and HDAC2 were measured before and after the intervention.

**Results:** Vitamin D supplementation resulted in a significant increase in both 25(OH)D_3_ and HDAC2 levels. Pulmonary function parameters showed an increasing trend, however, no statistically significant differences were observed.

**Conclusion:** Vitamin D supplementation was associated with increased HDAC2 levels, suggesting a potential anti-inflammatory effect. However, no significant improvement in pulmonary function was observed. Further studies are needed to determine its clinical impact.

## INTRODUCTION

Chronic Obstructive Pulmonary Disease (COPD) is a chronic progressive lung medical condition characterised by respiratory problems and chronic airflow blockage, often resulting from exposure to toxic chemicals, notably cigarettes. COPD is globally recognised as a leading cause of high mortality rates. This condition ranks as the third leading cause of death globally and has a prevalence of over 80% in nations with poor to moderate incomes. Within Indonesia, the estimated risk level is at 9.2 million individuals^4^. COPD involves persistent inflammation of the lungs, leading to a decline in quality of life by disrupting daily activities. COPD patients see incomplete recovery due to persistent inflammation resulting from the release of pro-inflammatory cytokines and chemokines. This inflammation can lead to tissue damage and obstruction of airflow in the lungs^2^. According to estimates, up to 85% of individuals with COPD are smokers due to the substantial reduction in lung function caused by exposure to tobacco smoke. Several specific medications, including Theophylline, long acting beta2-agonists (LABA), and inhaled corticosteroids, are beneficial in treating this condition. Nevertheless, the incidence of COPD cases continues to rise swiftly^8^.

Histone deacetylase 2 (HDAC2) is an enzyme essential for gene regulation in the process of chromatin remodelling. Individuals with COPD experience a reduction in HDAC2 gene expression, leading to elevated histone acetylation and the production of pro-inflammatory genes. The immunomodulatory action of vitamin D involves its binding to receptors, including macrophages, B cells, and T cells, therefore enhancing the immune response through the reduction of inflammation. At present, vitamin D is utilized as a medication that has a significant impact on cases of COPD due to its ability to influence associated systemic effects and its gene regulatory activities that are believed to provide protection against lung illness. Most of the synthesis of vitamin D occurs from pre-vitamin D3 in the skin of humans. By means of solar radiation, it will undergo synthesis from 7-dehydrocholesterol. Furthermore, fish liver oil, fish fat, liver, egg yolks, and cheese were identified as sources of vitamin D^8^.

According to Sultana et al., (2022), the presence of vitamin D inhibits the growth of Th 1 and Th17 cells, which play a role in inflammation. Consequently, insufficient levels of vitamin D can exacerbate symptoms by reducing lung function and increasing the frequency of lung exacerbations. The primary role of this vitamin is to facilitate bone metabolism. Furthermore, it has the ability to decrease the synthesis of inflammatory cytokines (IL-5, IL-9, and IL-13) in type-9 T-helper cells and so act as producers of the immune system. Nevertheless, this can be impaired if the patient suffers from a lack of vitamin D, which has implications for lung function, inflammation, and the prognosis of associated disorders^8^. Furthermore, there is a correlation between vitamin D insufficiency and chronic illnesses, including autoimmune disorders, cardiovascular disorders, and cancer^3^. Vitamin D, when used in long-term treatment and at the correct dosage, can effectively prevent acute exacerbations in individuals with COPD. Prolonged exposure to cigarette smoke as a catalyst for COPD will activate intracellular pathways (NF-B and MAPK p38) that are responsible for initiating inflammation. Furthermore, these results suggest that vitamin D may provide protection against inflammation and injury to lung tissue by blocking intracellular signaling pathways and stimulating profibrotic mediators^7^.

Prior research has demonstrated that vitamin D exerts a therapeutic impact by regulating HDAC2 levels in individuals with COPD. In order to decrease inflammation and enhance responsiveness to corticosteroids, vitamin D has the ability to enhance the activity of HDAC2. According to Janssens W et al. (2012), augmenting HDAC2 levels can mitigate oxidative damage caused by vitamin D supplementation. Additional research indicates a correlation between insufficient levels of vitamin D and pulmonary function, elucidated by variations in lung volume. This correlation can be linked to the association between COPD and the vitamin D status inside the body^13^. Furthermore, a similar study elucidated that the rising occurrence of vitamin D insufficiency among patients with COPD will lead to a decline in their quality of life. Therefore, the administration of vitamin D as a medication can effectively lower the occurrence of exacerbations. Nevertheless, there is currently no robust evidence indicating that this supplementation can effectively mitigate harm to the pulmonary organs^11^. The efficacy of vitamin D supplementation in managing COPD has been extensively established, however numerous studies have failed to yield consistent findings. Therefore, we undertook a study specifically targeting COPD patients. The objective of this study is to assess whether vitamin D supplementation can increase HDAC2 levels and has a potential to increase lung function in stable COPD patients.

## METHODS

### Research Design

This research was a pre-post test interventional study. The levels of HDAC2 were measured pre and post-supplementation of vitamin D given for 3 months. This study was conducted at the Pulmonary Clinic of Islamic Hospital Jemursari Surabaya from April to June 2024.

### Ethical Clearance

This study was approved by the Ethics Committee of *Komisi Etik Penelitian Kesehatan Rumah Sakit Islam Surabaya Jemursari*, with approval number 052/KEPK-RSISJS/V/2024. Written informed consent was obtained from all participants.

### Population

The samples in this study were COPD patients who meet the inclusion and exclusion criteria as follows: Inclusion criteria: 1). Diagnosed with COPD stage GOLD 2 and GOLD 3, 2). COPD patients who regularly seek treatment and receive standard bronchodilator therapy, 3). Willing to sign informed consent, 4). Patients with 25(OH)D levels < 32 ng/ml, 5). Male gender aged 40-80 years, 6). Normal liver function, 7). Normal kidney function. Exclusion criteria: 1). Suffering from respiratory diseases other than COPD, 2). Allergy to vitamin D. Drop-out criteria: 1). Moving to an address that cannot be contacted, 2). Passing away, 3). Not taking vitamin D for two consecutive weeks.

### Vitamin D Delivery

This vitamin exists in two active forms: vitamin D2 (ergocalciferol) and vitamin D3 (cholecalciferol). Calcitriol, also known as 1,25-(OH)2D3, is a pleiotropic hormone that emerges as the active form of vitamin D following its conversion by the kidneys from calcidiol, which is excreted by the liver. This conversion process is facilitated by the enzyme cytochrome P450 hydroxylase 1-alpha. Serum levels of 25(OH)D3 indicate vitamin D status. Patients received cholecalciferol at a dose of 5000 IU daily for three months. Patients who are chosen will thereafter be administered vitamin D at a dosage of 5000 IU for a duration of 3 months. Monthly monitoring of adherence and regular intake of vitamin D will be conducted. Upon completion of the study period, the patient’s blood will be collected for the purpose of assessing Vitamin D levels and conducting other test.

### Vitamin D level measurement

Serum levels of 25(OH)D3 were measured using a commercially available enzyme-linked immunosorbent assay (ELISA) kit (BT LAB, Cat. No. E1981 Hu), which is specifically designed for the quantitative determination of human 25(OH)D3 levels. The assay was performed according to the manufacturer’s instructions. In brief, serum samples were added to wells pre-coated with specific antibodies against 25(OH)D3, followed by the addition of enzyme-linked reagents to form antigen–antibody complexes. After incubation and washing steps to remove unbound substances, a substrate solution was added, producing a colorimetric reaction proportional to the concentration of 25(OH)D3 in the samples. The reaction was terminated using a stop solution, and absorbance was measured at 450 nm using a microplate reader.

### HDAC2 Examination

HDAC2 levels were measured using a commercially available enzyme-linked immunosorbent assay (ELISA) kit (BT LAB, Bioassay Technology Laboratory, Cat. No. E0988 Hu), according to the manufacturer’s instructions. The microplate was pre-coated with antibodies specific to human HDAC2. Samples were added to the wells, allowing HDAC2 present in the samples to bind to the immobilized antibodies. Subsequently, biotinylated anti-HDAC2 antibodies were added, followed by streptavidin–horseradish peroxidase (HRP), forming a sandwich complex. After incubation, unbound components were removed through washing. A substrate solution was then added, resulting in color development proportional to the concentration of HDAC2 in the samples. The reaction was terminated by the addition of a stop solution, and absorbance was measured at 450 nm using a microplate reader.

### Pulmonary Function Test

Pulmonary function tests were performed before and after Vitamin D supplementation using a calibrated spirometer to assess lung volumes and airflow. Standard spirometry parameters, including vital capacity (VC), forced vital capacity (FVC), forced expiratory volume in one second (FEV_1_), and forced expiratory flow at 25–75% of FVC (FEF25–75), were measured. Patients were instructed to abstain from smoking for at least 2 hours prior to the examination, avoid heavy meals before testing, and wear loose-fitting clothing. The use of bronchodilators was withheld prior to testing in accordance with standard recommendations (short-acting bronchodilators for at least 6–8 hours and long-acting bronchodilators for at least 12–24 hours). Anthropometric measurements, including height, were obtained to calculate predicted values based on reference standards from the Pneumobile Project Indonesia. All measurements were performed according to standardized procedures.

### Statistical Analysis

The statistical analysis was performed to evaluate differences in outcomes before and after the intervention. Continuous variables are presented as mean ± standard deviation (SD). The normality of the data distribution was assessed prior to analysis. Differences between pre- and post-intervention measurements, including serum Vitamin D levels, HDAC2 levels, and pulmonary function parameters, were analyzed using the paired t-test. A *p-value* of less than 0.05 was considered statistically significant. All statistical analyses were conducted using IBM SPSS Statistics.

## RESULTS

### Vitamin D levels

Vitamin D supplementation at a dose of 5000 IU for 3 months resulted in a significant increase in serum 25(OH)D3 levels compared to baseline (7.63 ± 2.73 ng/mL vs 14.26 ± 2.43 ng/mL, *p* = 0.023).

### Plasma HDAC2 levels

A significant increase in plasma HDAC2 levels was observed following Vitamin D supplementation (1.50 ± 1.02 ng/mL vs 3.90 ± 1.39 ng/mL, *p* = 0.006).

### Pulmonary Function Tests

Pulmonary function parameters, including vital capacity (VC), forced vital capacity (FVC), forced expiratory volume in one second (FEV_1_), and FEV_1_/FVC ratio, showed an increasing trend following Vitamin D supplementation. However, none of these changes reached statistical significance (all *p* > 0.05).

**Table 1.**
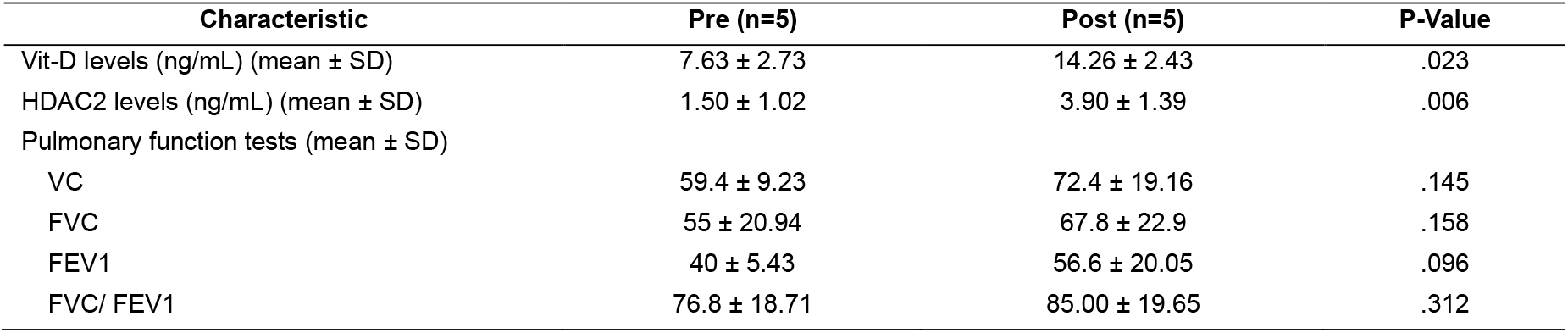
Lung function, Vitamin D and HDAC2 level before and after vitamin D 5000iu treatment.

**Figure 1.**
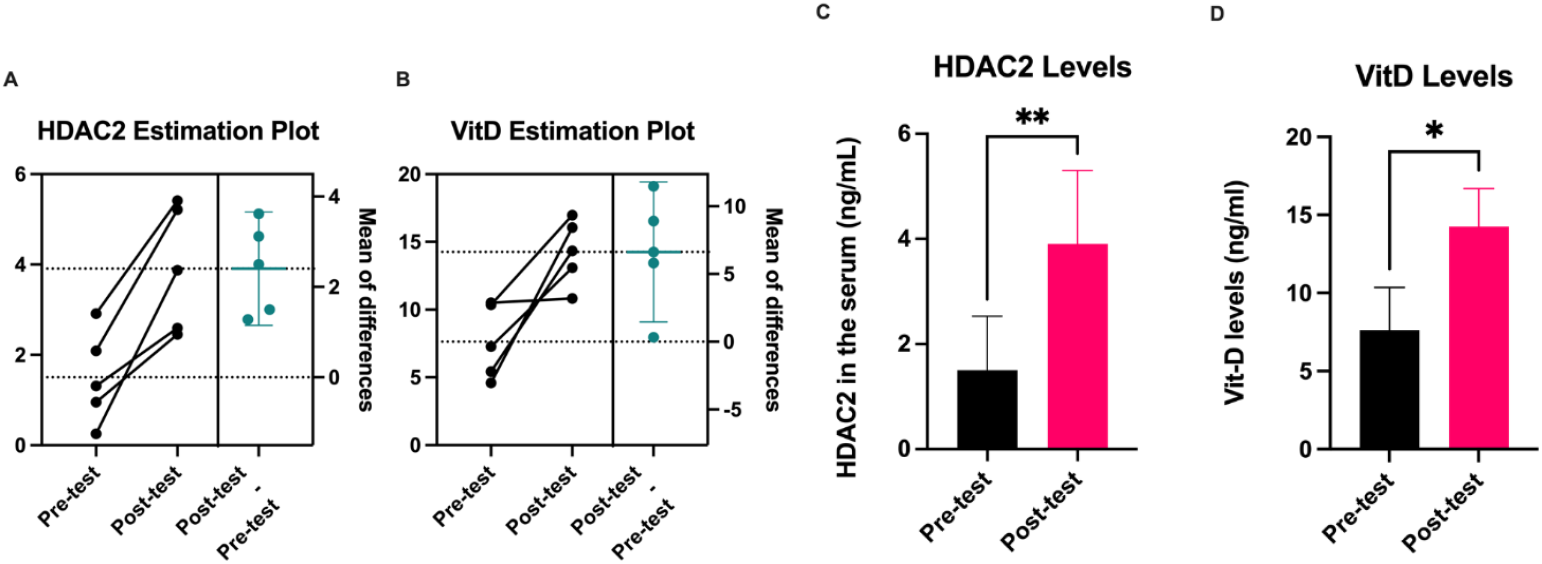
Vitamin D 5000iu effect in Vitamin D and HDAC2 serum levels after 3 months administration. A) Increased HDAC2 level. B) Increased Vitamin D level. C) Significant HDAC2 different pre-post. D) Significant Vitamin D different pre-post. Data represented as mean ± SEM. * p < 0.05, **p<0.01

## DISCUSSION

In this study, 25(OH)D levels increased from 7.63 ± 2.73 ng/mL prior to supplementation to 14.26 ± 2.43 ng/mL following Vitamin D supplementation. In elderly patients, daily supplementation with a Vitamin D dose of 5000 IU has been demonstrated to elevate 25(OH)D levels to over 85 nmol/L after three months. This level is deemed adequate to maintain the optimal function of Vitamin D in the body^10^. In the meta-analysis conducted by Li et al., (2020), the frequency of exacerbations in patients with COPD was significantly reduced by Vitamin D supplementation, particularly in those with severe Vitamin D deficiency and low Vitamin D levels (<25 nmol/L)^5^.

Vitamin D plays an important role in COPD due to its anti-inflammatory and immunomodulatory effects. Vitamin D inhibits the NF-κB signalling pathway, which is involved in excessive inflammatory processes, due to its anti-inflammatory properties. Decreased respiratory function, increased exacerbations, and hospitalisations are all linked to vitamin D deficiency. Furthermore, Vitamin D increases the expression of VDR (Vitamin D Receptor) in type II alveolar epithelial cells, which promotes surfactant synthesis and mitigates oxidative stress in the airways^8^.

In patients with COPD, HDAC2 is a critical regulator of inflammation. By suppressing NF-κB activity through histone deacetylation, HDAC2 reduces the activity of inflammatory genes, thereby inhibiting the expression of pro-inflammatory genes. The reduction of HDAC2 in COPD is linked to increased oxidative stress and cigarette smoke exposure, which speed up the degradation of HDAC2 by phosphorylating and nitrosylating specific residues. As corticosteroids are the primary treatment for COPD management, this condition exacerbates the patient’s response to them^15^. HDAC2 levels significantly increased following Vitamin D supplementation as indicated by this study. Although an increasing trend in pulmonary function parameters was observed following Vitamin D supplementation, no statistically significant improvement was detected. This may suggest that while Vitamin D exerts measurable effects at the molecular level, as reflected by increased HDAC2 levels, its impact on clinical outcomes such as lung function may require a longer duration of intervention or a larger sample size to become evident.

This study has several limitations that should be considered when interpreting the findings. First, the small sample size (n = 5) may limit the statistical power of the study and reduce the generalizability of the results. Second, the absence of a control group restricts the ability to establish a causal relationship between Vitamin D supplementation and the observed changes in HDAC2 levels and pulmonary function. Third, the relatively short duration of the intervention (three months) may not be sufficient to detect significant changes in clinical outcomes, particularly lung function parameters. Additionally, potential confounding factors such as variations in baseline disease severity, adherence to therapy, and environmental influences were not fully controlled. Therefore, further studies with larger sample sizes, longer follow-up periods, and controlled study designs are needed to confirm these findings.

## CONCLUSION

Vitamin D supplementation was associated with a significant increase in HDAC2 levels in patients with stable COPD, suggesting a potential anti-inflammatory effect at the molecular level. However, no statistically significant improvement in pulmonary function was observed. These findings indicate that Vitamin D may play a role in modulating inflammation in COPD, although its clinical impact requires further investigation in larger and well-controlled studies.

## DATA AVAILABILITY

The data supporting the findings of this study are available from the corresponding author upon reasonable request. All relevant data generated or analyzed during this study are included in this published article.

## CONFLICT OF INTEREST

The authors declare that there are no conflicts of interest regarding the publication of this study.

## FUNDING STATEMENT

This research was supported by Universitas Nahdlatul Ulama Surabaya (UNUSA) under the Letter of Assignment No. 023.92/UNUSA-LPPM/Adm.E/ST-Pen/III/2024.

## AUTHOR CONTRIBUTIONS

All authors contributed substantially to this study. Conceptualization and study design were performed by the authors. Data collection and analysis were conducted collaboratively. The manuscript was drafted and critically revised by all authors, and all authors approved the final version for publication.

## Notes

### Competing Interest Statement

The authors have declared no competing interest.

### Clinical Trial

TCTR20260405002

### Author Declarations

This study was approved by the Ethics Committee of Komisi Etik Penelitian Kesehatan Rumah Sakit Islam Surabaya Jemursari, with approval number 052/KEPK-RSISJS/V/2024.

## REFERENCES

1. Camargo, C. A., Toop, L., Sluyter, J., Lawes, C. M. M., Waayer, D., Khaw, K. T., et al. (2021). Effect of monthly vitamin d supplementation on preventing exacerbations of asthma or chronic obstructive pulmonary disease in older adults: Post hoc analysis of a randomized controlled trial. Nutrients, 13(2), 1–12. 10.3390/nu13020521

2. Global Initiative for Chronic Obstructive Lung Disease (GOLD). (2021). Global Strategy for the Diagnosis, Management, and Prevention of COPD.

3. Janssens, W., Lehouck, A., Decramer, M., & Gayan-Ramirez, G. (2012). Vitamin D and Chronic Obstructive Pulmonary Disease. In Vitamin D and the Lung (pp. 239–260). Humana Press. 10.1007/978-1-61779-888-7_11

4. Jarhyan, P., Hutchinson, A., Khaw, D., Prabhakaran, D., & Mohan, S. (2022). Prevalence of chronic obstructive pulmonary disease and chronic bronchitis in eight countries: a systematic review and meta-analysis. Bulletin of the World Health Organization, 100(03), 216–230. 10.2471/BLT.21.286870

5. Jolliffe, D.A., Greenberg, L., Hooper, R.L., Mathyssen, C., Rafiq, R., de Jongh, R.T., et al. (2019). Vitamin D to prevent exacerbations of COPD: systematic review and meta-analysis of individual participant data from randomised controlled trials. Thorax, 74(4), pp.337–345. 10.1136/thoraxjnl-2018-212092

6. Kerkhof, M., Voorham, J., Dorinsky, P., Cabrera, C., Darken, P., Kocks, J. W. H., et al. (2020). Association between COPD exacerbations and lung function decline during maintenance therapy. Thorax, 75(9), 744–753. 10.1136/thoraxjnl-2019-214457

7. Khan, D. M., Ullah, A., Randhawa, F. A., Iqtadar, S., Butt, N. F., & Waheed, K. (2017). Role of Vitamin D in reducing number of acute exacerbations in Chronic Obstructive Pulmonary Disease (COPD) patients. Pakistan Journal of Medical Sciences, 33(3). 10.12669/pjms.333.12397

8. Li, X., He, J., Yu, M., & Sun, J. (2020). The efficacy of vitamin D therapy for patients with COPD: a meta-analysis of randomized controlled trials. Annals of Palliative Medicine, 9(2), 286–297. 10.21037/apm.2020.02.26

9. Liao, W., Lim, A.Y., Tan, W.D., Abisheganaden, J. and Wong, W.F. (2020). Restoration of HDAC2 and Nrf2 by andrographolide overcomes corticosteroid resistance in chronic obstructive pulmonary disease. British Journal of Pharmacology, 177(16), pp.3662–3673. 10.1111/bph.15080

10. Mastaglia, S., Mautalen, C., Parisi, M., & Oliveri, B. (2006). Vitamin D2 dose required to rapidly increase 25OHD levels in osteoporotic women. European Journal of Clinical Nutrition, 60, pp. 681–687. 10.1038/sj.ejcn.1602369.

11. Mullin, M. L. L., & Milne, S. (2023). Vitamin D deficiency in chronic obstructive pulmonary disease. Current Opinion in Pulmonary Medicine, 29(2), 96–103. 10.1097/MCP.0000000000000935

12. Navarro, A. M. H.;, Hermosa, J. L. R., Rubio, M. C., Luis ílvarez-Sala, J., Vargas Centanaro, G., María, A., et al. (2022). Citation: Calle Rubio Testing for Vitamin D in High-Risk COPD in Outpatient Clinics in Spain: Clinical Medicine Testing for Vitamin D in High-Risk COPD in Outpatient Clinics in Spain: A Cross-Sectional Analysis of the VITADEPOC Study. J. Clin. Med, 2022, 1347. 10.3390/jcm11051347

13. Nur Aini, V. F., Widajati, E., & Mustafa, A. (2023). Pengaruh Suplementasi Vitamin D terhadap Fungsi Paru pada Pasien Penyakit Paru Obstruktif Kronik (PPOK). NUTRITURE JOURNAL, 2(2), 78. 10.31290/nj.v2i2.3862

14. Sultana, N., Ali, T., Bennoor, K. S., Hossain, M. A., Mahmud, M. B., Hassan, S., et al. (2022). Vitamin D3 supplementation on lung functions and exercise tolerance in D3 deficient asthma COPD overlap syndrome patients - A randomized controlled trial. Journal of Bangladesh Society of Physiologist, 17(1), 21–33. 10.3329/jbsp.v17i1.63213

15. Zwinderman, M.R., de Weerd, S. and Dekker, F.J. (2019). Targeting HDAC complexes in asthma and COPD. Epigenomes, 3(3), p.19. 10.3390/epigenomes3030019

